# Bone health risk assessment in a clinical setting: an evaluation of a new screening tool for active populations

**DOI:** 10.1101/2020.08.07.20170142

**Authors:** Nicola Keay, Gavin Francis, Karen Hind

## Abstract

**Introduction:** Risk factors for poor bone health are not restricted to older, sedentary populations for whom current screening is focused. Furthermore, access to dual X-ray absorptiometry scanning can be limited in clinical practice. The purpose of the current study was to develop a bone health-screening tool suitable for inclusion of both younger and active populations, combined with radiofrequency echographic multi spectrometry technology (REMS).

**Methodology:** 88 participants attending a physiotherapy clinic in the UK were recruited to the study: 71 women (mean age 41.5 SD 14.0 years); 17 men (mean age 40.2 SD 14.9 years). Participants completed an online bone health-screening questionnaire developed specifically for this study covering a range of lifestyle, physiological factors, combined with medical interview and received bone mineral density (BMD) measurement at the lumbar spine and femoral neck using REMS.

**Results:** Scoring of the bone health-screening questionnaire produced a distribution of bone health scores, with lower scores suggesting a higher risk for poor bone health. In women, scores ranged from -10 to +12, mean score 2.2 (SD 4.8). In men, scores ranged from 0 to 12, mean score 6.9 (SD 3.2). A positive correlation was observed between the bone health score derived from the questionnaire and lumbar spine and femoral neck BMD Z-scores (p<0.01).

**Conclusions:** This new and comprehensive bone health-screening questionnaire with interview was effective in identifying active individuals at risk of bone fragility, who might be missed by current screening methods. The use of REMS technology to measure bone health, was feasible in the clinical setting.

## Introduction

Deficits in skeletal health can arise as part of the ageing process, for example during and after menopause in women (^1^). Lifestyle factors can also contribute to impaired bone health, such as a lack of weight bearing exercise (^2^). Conversely, high exercise training loads can result in a low energy availability state and the clinical consequences of relative energy deficiency in sport (RED-S) (^3^), which include adverse effects on bone health (^7^). Suboptimal bone health increases the risk of bone stress injuries in these exercisers, including stress fractures (^4^) and complete fractures (^5^). These types of bone injuries have consequences in terms of morbidity and athletic performance, as well as mortality in the older population (^1^). Therefore, early identification of those with impaired bone health is crucial. The Fracture Risk Assessment Tool FRAX^®^ questionnaire is an effective tool for the identification of adults aged >40 years, at high risk of fragility fracture (^6^). In exercising populations, a validated screening questionnaire has been developed for male cyclists at risk of low bone density (^7^) but there is currently no screening tool for the active, general population.

Bone mineral density (BMD) is a marker of bone strength and is commonly measured using dual-energy X-ray absorptiometry (DXA) in accordance with the World Health Organisation (WHO) guidelines for the assessment of osteoporosis (^8^) and can be combined with the clinical information collected in the FRAX^®^ questionnaire. It has been suggested by the WHO that further areas of research are required to inform on risk prediction, including considering other risk factors and methods of assessing poor bone health(^8^). There is also a lack of screening tools for populations younger than 40 years and for active populations, who may be at risk of impaired bone health. Radiofrequency Echographic Multi Spectrometry (R.E.M.S.) (^9^) technology has recently been introduced for the assessment of bone strength in adults, and unlike DXA, does not involve ionising radiation, making it particularly useful for screening purposes. In clinical validation studies, REMS has been shown to have good level of accuracy and precision, in addition to significant agreement with DXA in a multicentre trial involving 1,914 females (^10^). From this trial, the REMS intra-operator precision, expressed as root mean-square coefficient of variation (RMS-CV) was 0.38% (95% confidence interval: 0.28–0.48%) for lumbar spine and 0.32% (0.24– 0.40%) for femoral neck. The corresponding least significant change for the 95% confidence level, calculated via the ISCD precision calculator (available at http://www.iscd.org/resources/calculators/), was 1.05% for lumbar spine and 0.88% for femoral neck (^11^).

The objective of the current study was to explore the effectiveness of a new bone health-screening tool with clinical interview, developed for the identification of active individuals at risk of poor bone health. Together with evaluation of the practicality of REMS for bone strength assessment in this population, in the clinical setting.

## Materials and Methods

### Participants

Participants (n=88) were recruited from clients attending a private physiotherapy clinic in Bath, United Kingdom. The physiotherapy clinic provides physiotherapy, strength and conditioning programmes and clinical input for a range of conditions, including those exercisers with suspected low energy availability. Invitation for participants was also disseminated through contacts in the vicinity such as university, sport clubs and healthcare providers referring to the physiotherapy practice. The inclusion criteria were males and females over the age of 20. The study was approved by the university research ethics committee and all participants provided informed consent prior to taking part.

### Bone Health Screening Questionnaire

All participants completed the bone health questionnaire (supplementary file 1). The questionnaire was specifically designed to quantify an overall clinical assessment of bone health, taking into account recognised risk and mitigating factors for bone health from published evidence to date. This included the risk factors assessed in the FRAX^®^ clinical assessment questionnaire (^12^): body mass index (BMI), medical history (history of fractures, treatment with steroids, rheumatoid arthritis) and lifestyle factors (smoking, alcohol intake). In addition, our bone health-screening questionnaire included questions on exercise levels, dietary habits and indicators of sex steroid hormonal function. The detail included weekly skeletal loading and non-skeletal loading exercise levels; dietary habits, diagnosed eating disorders, relative energy deficit in sports (RED-S). In women, questions included current menstrual status and menstrual history; in men, average number of weekly morning erections to indicate testosterone levels (^13^). Subjective reports of sleep quality and fatigue/freshness, alongside bone injury history and fundamental medical background were also gathered. Clinical medical interview was conducted to verify and clarify any areas in the questionnaire.

Participant responses were scored as shown in supplementary file 2. This scoring system is similar to systems published for validated screening questionnaires elsewhere including FRAX^®^ and clinical assessment tools for active females (^14^), exercising men (^3^) and for exercising men and women (^15^).

### Bone strength assessment

Each participant received lumbar spine and hip BMD scans using R.E.M.S. technology with an Echolight S machine (Echolight, Certification Mark MED31204, Italy). For each scan, participants were positioned supine on a treatment couch. Initially the lumbar spine was scanned followed by the hip and ultrasound gel was distributed over the region to be scanned. A locating scan was performed to set optimal transducer focus and scan depth, in order to visualize the target bone interphase in the central part of the echographic field of view, immediately below the focus position. After this locating scan, an acquisition scan was performed for the diagnostic evaluation of the regions of interest. All scans were performed by a medical doctor and a research nurse who had both successfully completed the Echolight Clinical Course and attained practical experience in using the technology. Before each scanning session, a quality control check was performed, using the phantom provided by the manufacturer.

### Statistical analysis

All data were assessed using the python programming language software Pandas version 1.0.3 on Zenodo and Statsmodels by Seabold, Skipper, and Josef Perktold. Data were interrogated for relationships between bone health score and BMD measured at the lumbar spine and femoral neck. Significance was assessed using a T-test, applied to the linear regression coefficient between continuous variables, and an F-test on the correlation coefficient, with a threshold p-value of 0.05.

### Practicality evaluation

Clinical medical explanations were provided by the medical doctor for each participant verbally and in written form, together with printout of the scan charts. This included explanation of the charts and relating findings to information reported in the questionnaire and interview. From this discussed review, personalised lifestyle and medical educational based information was delivered. Any feedback from participants on the experience of the scan procedure and usefulness of information relating to the appointment was noted. For example, if the scan was a confortable experience and whether the information provided was helpful in terms of informing and guiding any actionable, practical steps to take.

## Results

A total of 88 participants completed the bone health questionnaire and received lumbar spine and femoral neck BMD assessment by REMS. The participants comprised women n=71, mean age 41.5, SD 14.0 (range 20 to 71 years), mean BMI 21.7 SD 4.0, where 22 were postmenopausal; and men n=17, mean age 40.2, SD 14.9 (range 22 to 70 years); mean BMI 24.2 SD 3.7.

Scoring of the bone health-screening questionnaire and interview produced a distribution of scores shown in Figure 1. Bone health scores for women ranged from -10 to +12, with a mean of 2.2 (SD 4.8), and men’s scores ranged from 0 to 12 with a mean of 6.9 (SD 3.2). Where negative scores indicated potential risk to bone health and positive values indicated potential positive factors.

**Figure 1.**
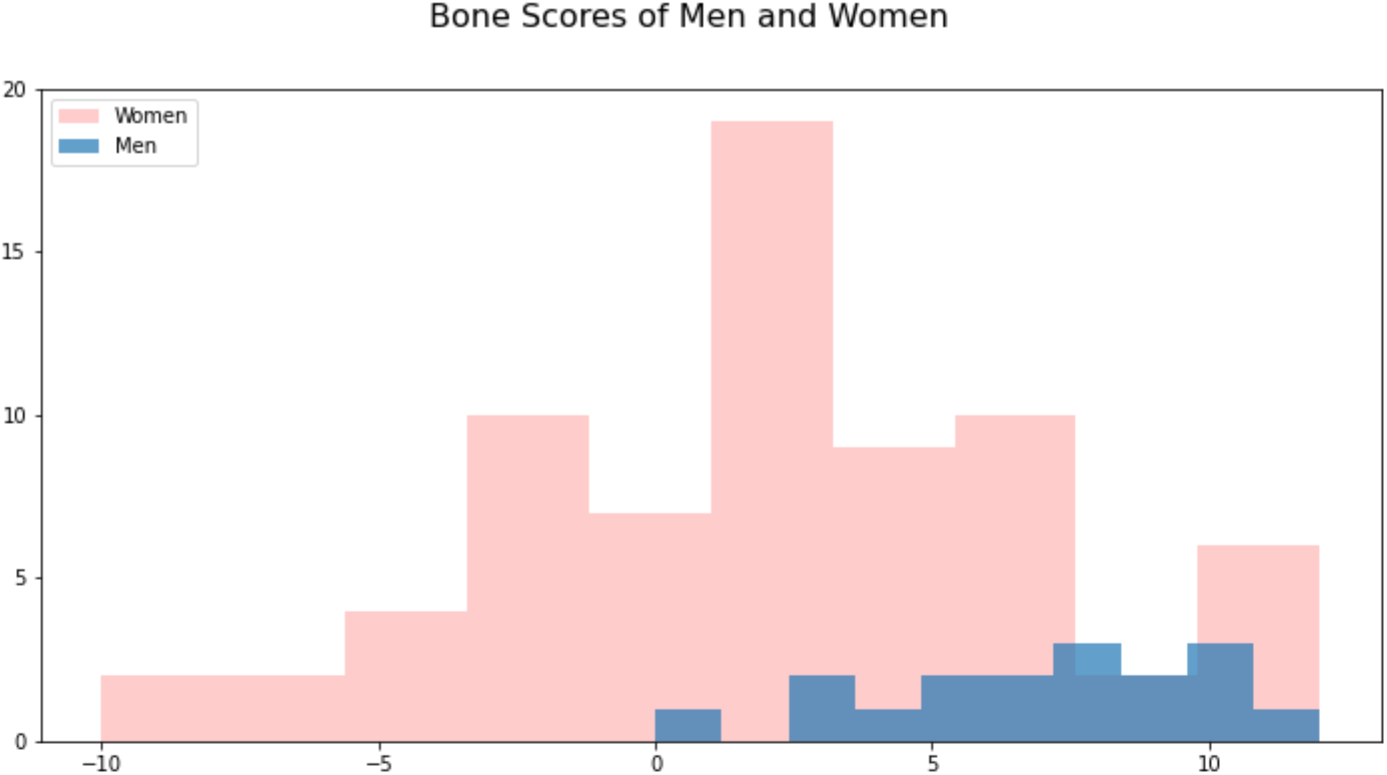
Bone Heath Scores calculated from screening questionnaire

Findings from REMS: 19 participants were fund to have lumbar spine BMD Z-score <-1.0 which is the criteria for indication of bone health consequences in RED-S clinical assessment tool (^15^)

Figure 2 shows the relationship between bone health score and lumbar spine BMD Z-score for all participants. A positive correlation was observed between the bone health score derived from the questionnaire and lumbar spine BMD Z-score (R=0.14, p<0.01), indicating that higher bone health scores were associated with better lumbar spine BMD.

**Figure 2.**
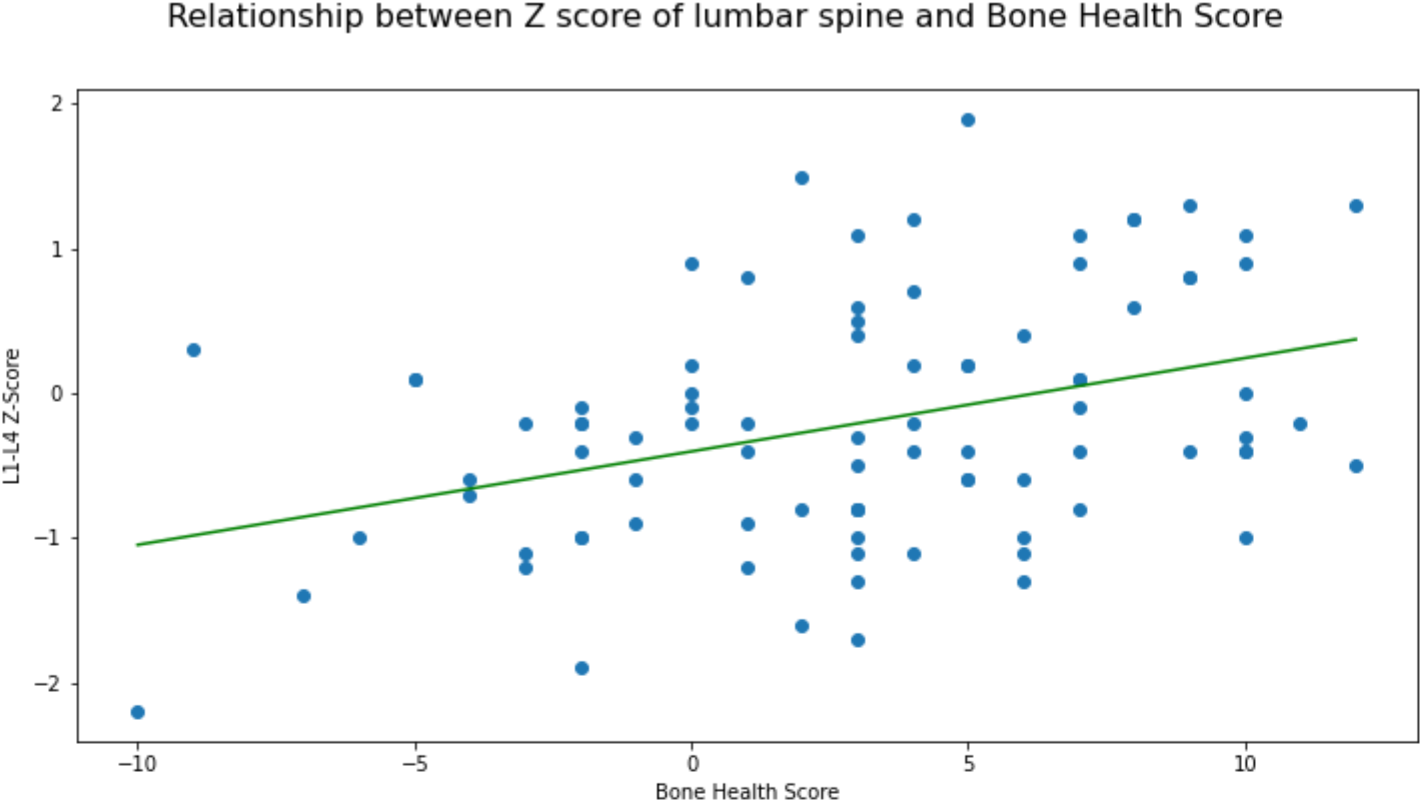
Graph of lumbar spine bone mineral density Z-score against bone health scores

Figure 3 shows a similar association between bone health score and femoral neck BMD Z-score (R = 0.27, p<0.01).

**Figure 3.**
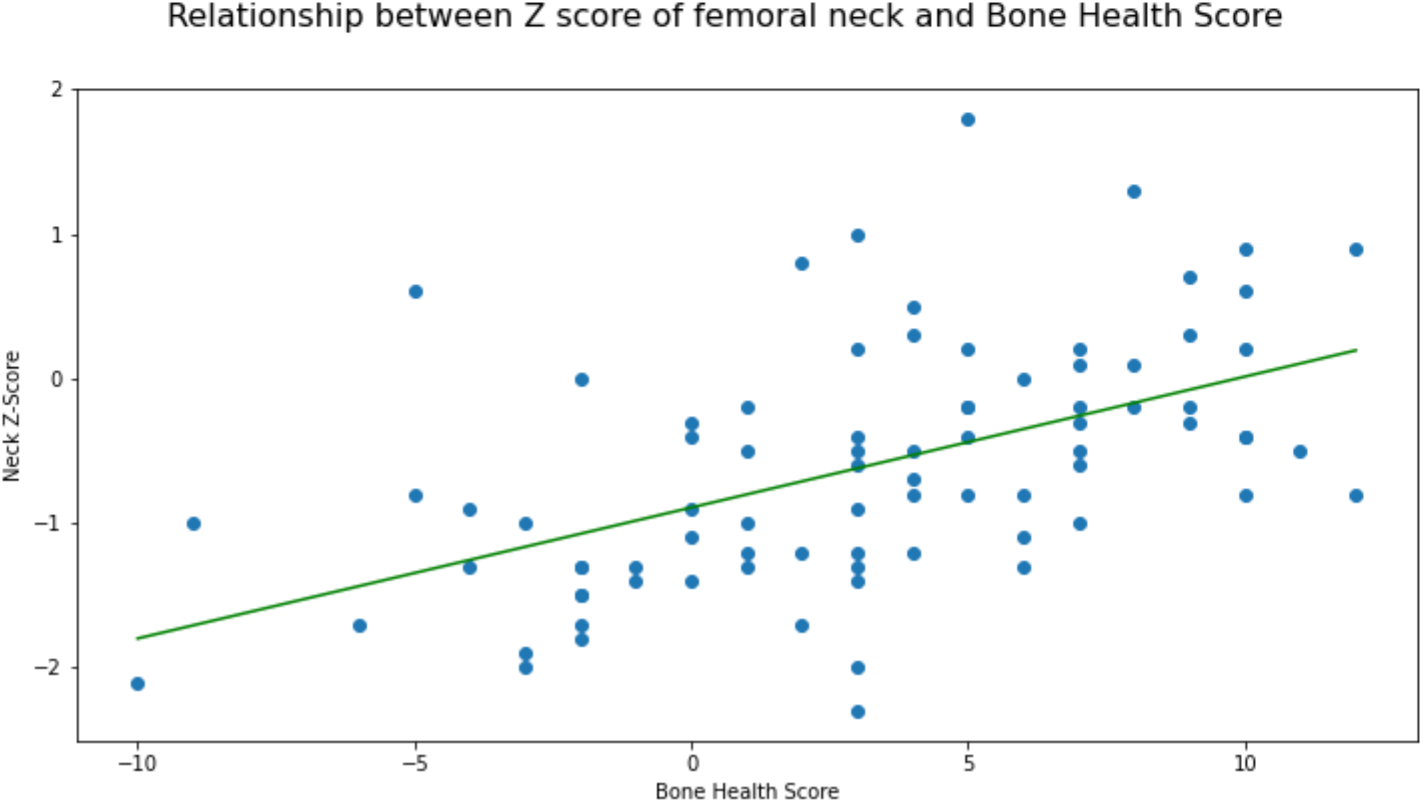
Graph of femoral neck bone mineral density Z-score against bone health scores

Practicality evaluation of this form of bone health assessment from participant feedback was favourable; in that the process of completing questionnaire and scan was efficient. Immediate explanation of results and sign posting was indicated as being helpful. 30 minutes per participant was allocated for scanning and feedback of results, which was found to be practical, provided breaks after 6 successive participants were added. For the healthcare professionals with previous “hands on” clinical experience, scanning with the REMS device was technically relatively easy to master. As indicated in earlier clinical evaluation studies, operator training is important to reduce acquisition errors (^10^). From this study, which included young, active participants, it was noted that where low energy availability was suspected, an increased gas in the abdomen could sometimes make locating lumbar spine challenging.

## Discussion

In this study we have demonstrated the potential of a new bone health-screening questionnaire for the identification of active individuals at risk of low bone density. This new questionnaire is not restricted to those over 40 years, rather includes younger adults potentially at risk of suboptimal bone health and takes into account a range of lifestyle factors. REMS was found to be potentially useful scanning modality in the clinical setting.

The FRAX^®^ clinical tool is a well-established questionnaire to assess the risk of fragility fracture (osteoporotic fracture). However, the focus of questions is on underlying medical conditions and the limited lifestyle factors of smoking and drinking alcohol that predispose to low BMD. Furthermore this questionnaire is only applicable to those over 40 years of age (^6^). Although extra information can be added through results from DXA, this type of scan is not readily available or accessible for those under 40 years of age. Nevertheless, younger, active populations can also be at risk of impaired bone health, in particular exercisers, where low energy availability can arise unintentionally or intentionally (^3^). In the case of intentional low energy availability, this includes exercisers on a spectrum that can range from disordered eating to a clinically diagnosed eating disorder (^16^). In this way, even if eligible for completing FRAX questionnaire from an age point of view, the risk of poor bone health could be missed. In terms of medical conditions, as those in low energy availability may not have an eating disorder meeting diagnostic criteria. Furthermore, there is evidence that BMI can still be within the normal range in a low energy availability state, explained at least partly, by a compensatory alteration in metabolic rate (^17^). Additionally, no negative lifestyle factors for bone health would be reported for these exercisers in terms of drinking large amounts of alcohol or smoking.

Although healthy lifestyle behaviours, including weight-bearing exercise are well recognised to have a beneficial osteogenic effect (^18^), high exercise training loading loads that are not matched by sufficient dietary intake, can lead to the adverse health and performance consequences of low energy availability, described in the clinical syndrome of RED-S (^16^). Significantly low energy availability can impact both male and female exercisers and bone stress injuries are a well-documented outcome (^4^). In terms of screening for low energy availability as a risk factor for poor bone health in athletes, the low energy availability female questionnaire (LEAF-Q) is a validated tool (^14^) with emphasis on menstrual history and status. For men, application of a sport-specific energy availability questionnaire combined with clinical interview (SEAQ-I) proved effective in identifying those male cyclists at risk of the bone health consequences of RED-S (^7^). In the current questionnaire, a greater range of risk factors was considered. In particular, men were asked to respond to a question assessing testosterone levels through number of morning erections per week, as sex steroids in both male and females are validated indicators of low energy availability and the outcome of bone stress injuries in exercisers (^4^). In this way the bone health-screening questionnaire in this study can be administered to both men and women, employing validated scoring systems. The sample included significantly more female participants, possibly reflecting a greater interest and awareness of bone health issues amongst women.

Furthermore, there is evidence that poor bone health as a consequence of RED-S is not a condition that is limited to elite athletes competing in sport. Rather this syndrome can occur in dancers, aspiring amateur athletes and those who are not involved in competitive sport (^19^), but who are exercise-dependent (^20^) and this is recognised as an indicator of disordered eating and hence low energy availability and poor bone health (^3^). This situation can potentially be more likely to occur during the lock down during pandemics, where exercisers, dancers and athletes might look to increase usual exercise training to alleviate stress levels (^21^).

Promptly identifying exercisers at risk of poor bone health is crucial. This is demonstrated by the finding that bone deficits developed due to low energy availability may not be fully reversible, even once menstrual function is restored in female exercisers (^22^). Furthermore, the consequences of impaired peak bone mass accumulation due to high training loads from a young age (^23^) might not be evident till later in life (^**Error! Bookmark not defined**.^) and it has been suggested that accumulation of peak bone mass is more significant than age of menopause, in the development of osteoporosis (^24^).

The bone health-screening questionnaire developed and applied in this study has the potential to identify younger, active populations at risk of suboptimal bone health, who would not be identified by FRAX, nor necessarily by questionnaires targeting high-level athletes. Once identified, preventative interventions can be put in place, which include lifestyle changes and pharmacological treatment, in selected cases, to prevent progression and in some cases improve bone health. Evidence for positive outcomes of these behavioural interventions are reported in studies of the older population (^25^) and male cyclists (^26^). In terms of pharmacological interventions in younger active female populations, for those with functional hypothalamic amenorrhoea, hormone replacement therapy has been shown to more bone protective than no treatment or combined oral contraceptive pill (^27^). Therefore this bone health screening questionnaire, applicable to all active age groups, provides a cost effective, practical clinical tool to identify those at risk of poor bone health, who would not be eligible or could be missed using current the bone health screening. Once identified as being at risk, bone health imaging and targeted preventative strategies could be implemented and changes in bone health monitored.

A significant correlation was demonstrated between bone health score and lumbar spine BMD Z-score, a trabecular rich skeletal site and femoral neck BMD Z-score, as assessed by REMS technology. Although relatively new to the field, REMS shows promise for the diagnosis of impaired bone health and initial studies indicate potential for fracture prediction (^28^, ^29^). It is recommended that future studies extend the effectiveness of the bone health-screening questionnaire against DXA and for the prediction of fracture. None-the-less, as a non-ionising radiation approach to the assessment of bone health, REMS offers potential for longitudinal monitoring, particularly in younger populations.

## Limitations and further work

This study attracted a larger number of female participants than male, further investigation to actively recruit more male participants would be informative. Whilst REMS technology already shows great promise, DXA is ultimately the current “gold standard” for assessing BMD. Further work validating the bone health questionnaire for young active populations against BMD from DXA will be the next step.

## Conclusions

This study indicates that a comprehensive bone health-screening questionnaire, applicable to active men and women of all ages, is a practical and cost-effective clinical tool. REMS technology was found to be an accessible, feasible method of assessing bone health. Further assessment of both the questionnaire and REMS scanning, for example with DXA and future fracture incidence, will be valuable in extending the ability to identify and monitor those at risk of suboptimal bone health.

## Data Availability

The data that support the findings of this study are available from the corresponding author upon reasonable request.

## Acknowledgements

Thank you to Bonosso Ltd, UK distributor of Echolight S.p.A, for providing loan of the Echolight scanner.

## Illustration legends

Supplementary file 1 Bone Health Screening Questionnaire

Supplementary file 2 Bone Health Screening Questionnaire scoring system

Figure 1 Bone Heath Scores calculated from screening questionnaire

Figure 2 Graph of lumbar spine bone mineral density Z-score against bone health scores

Figure 3 Graph of femoral neck bone mineral density Z-score against bone health scores

## Supplementary file 1 Bone Health Screening Questionnaire

Name

DOB

Age

Contact email and mobile

If you have had DXA scan, please state

Lumbar spine (total L1-4) Z score

T score Femoral neck Z score T score

### Background Information

Height

Weight

1. BMI (calculated)
2. Do you have any medical conditions?
3. Please list any prescribed medications you are taking
4. Please list any supplements you are taking
5. Do you/have you ever smoked?
6. How would you rate your sleep quality? Scale 0-5 where: 0=poor and 5= good sleep
7. How would you rate freshness/fatigue? Scale 0-5 where: 0=very fatigued and 0=totally fresh

### Exercise

8. How many hours do you exercise per week?
  a. Weight bearing exercise (eg running, dancing)
  b. Resistance exercise (eg S&C, Pilates, Yoga)
  c. Non weight bearing (eg swimming, cycling)

### Injures

9. If you have had any fractures, please list, stating site of fracture

### Nutrition

10. Are you vegetarian?
11. Are you vegan?
12. Have you ever been diagnosed as having RED-S (relative energy deficiency in sport)?
13. Have you ever had an eating disorder?
14. How many portions of dairy do you have per week on average? (where portion= carton yogurt, glass of milk (in tea/coffee/cereal), cheese serving etc)
15. How many cups of caffeinated coffee do you drink per day?
16. How many units of alcohol do you drink per week?

### Females

17. What age did your periods start?
18. Are you periods regular? (excluding withdrawal bleeds on combined oral contraceptive pill)
  - No, I am using hormonal contraception
  - More than 9 periods per year
  - Less than 9 periods per year
  - No periods for more than 6 months
19. If you have experienced 6 months or more without periods, for how many years was this the situation?
20. Are you on hormonal contraception? Please state what
21. What age did you reach menopause (if relevant)?
22. If you are menopausal, do you take HRT? Please name

### Males

23. To assess testosterone status, please state number of morning erections per week (on average) 0 to 7

## Supplementary file 2 Scoring Questionnaire for Bone Health

Height

Weight

1. **BMI** (calculated) **<20 score -1** **<18 score -2**
2. Do you have any medical conditions? **-2 any eating disorder, FHA** **-1 amenorrhoea (not ED), DM, hyperthyroidism** **-1 RA, coeliac** **+1 None**
3. Please list any prescribed medications you are taking **-2 steroids** **-1 NSAID, PPI** **+1 None**
4. Please list any supplements you are taking **Yes: Vit D = +1**
5. Do you/have you ever smoked? **-1 yes** **+1 no**
6. How would you rate your sleep quality? 0-5 where 0=poor and 5= good sleep **-1 for 0** **0 for 3** **+1 for 5**
7. How would you rate freshness/fatigue? 0-5 where 0=very fatigued and 0=totally fresh **-1 for 0** **0 for 3** **+1 for 5**

### Exercise

8. How many hours do you exercise per week?
  a. Weight bearing exercise (eg running, dancing) **0 hrs = -1; 1-2 hrs = 0; 3-15 hrs = +1; >15 hrs = -1 score**
  b. Resistance exercise (eg S&C, Pilates, Yoga) **0 hrs = 0; >0hrs = +1 score**
  c. Non weight bearing (eg swimming, cycling) **score = 0**

### Injures

9. If you have had any fractures, please list, stating site of fracture **-3 trabecular rich (spine, sacrum, pelvis, calcaneus)** **-2 other sites (long bones eg legs, NOF)** **-1 trauma #**

### Nutrition

10. Are you vegetarian? **-1 yes +1 no**
11. Are you vegan? **-2 yes, +1 no**
12. Have you ever been diagnosed as having RED-S (relative energy deficiency in sport)? **Yes = -2, No = +1 (if already counted above under medical condition = 0)**
13. Have you ever had an eating disorder? **Yes = -2, No = +1 (if already counted above under medical condition = 0)**
14. How many portions of dairy do you have per week on average? (where portion= carton yogurt, glass of milk (in tea/coffee/cereal), cheese serving etc) **<7 = -1**
15. How many cups of caffeinated coffee do you drink per day? **>4 portions per day = -1**
16. How many units of alcohol do you drink per week? As per FRAX^**®**^ **>3 units per day (21 units per week) = -1**

### Females

17. What age did your periods start? **>16 years = -2**, **Otherwise +1**
18. Are you periods regular? (excluding withdrawal bleeds on combined oral contraceptive pill) **>9 per year (eumenorrhoea)= +1** **pregnancy, menopause, COCP 0** **<9 (oligomenorrhoea) = -2** **>6 months no periods (secondary amenorrhoea) = -3**
19. If you have experienced 6 months or more without periods, for how many years was this the situation?
20. Are you on hormonal contraception? Please state what
21. What age did you reach menopause (if relevant)? **<45 yrs = -2** **<51 yrs = -1** **>51 yrs = +1** **>55 yrs = +2**
22. If you are menopausal, do you take HRT? Please name **Yes = +1** **No = -1** **Males**
23. To assess testosterone status, please state number of morning erections per week (on average) 0 to 7 **<2 = -2** **<4 = -1** **>4 = +1**

